# HPV vaccination of girls in the German model region Saarland: Insurance data-based analysis and identification of starting points for improving vaccination rates

**DOI:** 10.1101/2021.10.01.21264397

**Authors:** Anna Marthaler, Barbara Berkó-Göttel, Jürgen Rissland, Jakob Schöpe, Emeline Taurian, Hanna Müller, Gero Weber, Stefan Lohse, Thomas Lamberty, Bernd Holleczek, Harry Stoffel, Gunter Hauptmann, Martin Giesen, Christiane Firk, Alexandra Schanzenbach, Florian Brandt, Heike Hohmann, Quirin Werthner, Dominik Selzer, Thorsten Lehr, Stefan Wagenpfeil, Sigrun Smola

**Affiliations:** Institute of Virology, Saarland University Medical Center, Kirrbergerstr. Building 47, D-66421 Homburg, Germany; Helmholtz Institute for Pharmaceutical Research Saarland (HIPS), Helmholtz Centre for Infection Research, Saarland University Campus, D-66123 Saarbrücken, Germany; Institute for Medical Biometry, Epidemiology and Medical Informatics, Saarland University Medical Center, Kirrbergerstr. Building 86, 66421 Homburg, Germany; Physical Geography and Environmental Research, Saarland University, Am Markt Zeile 2, 66125 Saarbrücken, Germany; Ministry of Health, Social Affairs, Women and the Family, Franz-Josef-Röder-Str. 23, 66119 Saarbrücken, Germany; Saarland Cancer Registry, Neugeländstraße 9, 66117 Saarbrücken, Germany; Kassenärztliche Vereinigung Saarland, Europaallee 7-9, 66113 Saarbrücken, Germany; AOK Rheinland-Pfalz/Saarland, Halbergstraße 1, 66121 Saarbrücken, Germany; IKK Südwest, Trierer Str. 4, 66111 Saarbrücken, Germany; Clinical Pharmacy, Saarland University, Saarland University Campus, D-66123 Saarbrücken, Germany

## Abstract

In Germany, the incidence of cervical cancer, a disease caused by human papillomaviruses (HPV), is higher than in neighboring European countries. HPV vaccination has been recommended for girls since 2007. However, it continues to be significantly less well received than other childhood vaccines, so its potential for cancer prevention is not fully realized.

To find new starting points for improving vaccination rates, we analyzed pseudonymized routine billing data from statutory health insurers in the PRÄZIS study in the federal state Saarland serving as a model region. We show that lowering the HPV vaccination age to 9 years led to more completed HPV vaccinations already in 2015. Since then, HPV vaccination rates and the proportion of 9-to 11-year-old girls among HPV-vaccinated females have steadily increased. However, HPV vaccination rates among 15-year-old girls in Saarland remained well below 50% in 2019. Pediatricians vaccinated the most girls overall, with a particularly high proportion at the recommended vaccination age of 9-14 years, while gynecologists provided more HPV catch-up vaccinations among 15-17-year-old girls, and general practitioners compensated for HPV vaccination in Saarland communities with fewer pediatricians or gynecologists. We also provide evidence for a significant association between attendance at the U11 or J1 medical check-ups and HPV vaccination. In particular, participation in HPV vaccination is high on the day of U11. However, obstacles are that U11 is currently not funded by all statutory health insurers and there is a lack of invitation procedures for both U11 and J1, resulting in significantly lower participation rates than for the earlier U8 or U9 screenings, which are conducted exclusively with invitations and reminders.

Based on our data, we propose to restructure U11 and J1 screening in Germany, with mandatory funding for U11 and organized invitations for HPV vaccination at U11 or J1 for both boys and girls.

## Introduction

Invasive cervical cancer is a consequence of persistent human high-risk papillomavirus (HPV) infection (1, 2). Cervical cancer remains the most important vaccine-preventable HPV-associated cancer, with 604,127 cases and 341,831 deaths in 2020 worldwide (3). In Germany, the incidence of cervical cancer in 2016 was 8.7 per 100,000 women (age-standardized, European population (4)), while neighboring countries such as Switzerland, Austria, France and the Netherlands have lower incidences (4, 5). For cancers of the vagina, vulva, penis, anus, and oropharynx, HPV-associated and HPV-independent forms of cancer are existing (6, 7). In recent years, there has been a particularly sharp increase in potentially HPV-associated oropharyngeal cancers in women in Germany (8, 9). The same is true for vulvar cancer (7, 10). In particular, the incidence of vulvar cancer in Germany is also relatively high compared to other European countries (4). With the exception of cervical cancer in women and HIV-positive men who have sex with men, who have a 100-fold increased risk of anal cancer, there is currently no organized screening in Germany for HPV-associated cancers (11-14).

HPV vaccination, with high immunogenicity particularly in preadolescents, high efficacy, and a good safety profile, is an effective measure for sustained reduction in the burden of HPV-associated cancers. Because HPV is sexually transmitted, vaccination is most efficacious when given prophylactically to HPV-naïve individuals, ideally before first sexual intercourse. Population studies from other countries with school-based vaccination such as Australia show high vaccination rates (>80%) providing herd immunity in the population beyond individual protection. This makes cervical cancer elimination a realistic goal (15). In Australia, the 4-valent and subsequent 9-valent vaccines have been used, covering the high-risk viruses HPV16, 18 (4-valent) or 16, 18, 31, 33, 45, 52, and 58 (9-valent), as well as the low-risk viruses HPV6 and 11. Already in 2013 a substantial reduction in cervical precancerous lesions was observed (4). A meta-analysis found that in countries with HPV vaccination rates >50% in girls, there was a 68% reduction in HPV16 and 18 infections as early as 2014 (16). More recent meta-analyses of studies between 2014 to 2018 showed a decrease in HPV16 and 18 infections by 83% 5-8 years after vaccination introduction in 13- and 19-year-old females, and of anogenital warts by 67% in 15-19-year-old females (17). Laryngeal papillomas also showed a significant decrease over the course of HPV vaccination (18). Herd immunity was reflected in an 81% and 51% reduction in genital warts in heterosexual men under 21 and 21-30 years of age, respectively (19). A reduction of high-grade cervical dysplasia (CIN2+) by 51% in young women aged 15-19 years and by 31% in 20-24 year-old females was observed (17). Most importantly, a Swedish registry study of girls and women aged 10-30 years conducted between 2006 and 2017 found a substantially lower risk of invasive cervical cancer at the population level (88% reduction after covariate adjustment) if HPV vaccination was administered before the age of 17 years (20). This indicates that timely HPV vaccination is not only able to prevent viral infection, benign warts and precancerous lesions but also cancer.

In Germany, the HPV vaccination for girls has been recommended by the Standing Committee on Vaccination (STIKO) in 2007 and has since been one of the mandatory benefits of statutory health insurance. Originally, the vaccination age was set at 12-17 years (21). In 2014, the vaccination age was lowered to 9-14 years and catch-up vaccination was recommended until the age of 17 years (22). In addition, the vaccination schedule was adjusted to the age of the vaccinated with 2 or 3 doses, respectively. In 2018, the HPV vaccination recommendation was extended to boys (23), as about 6,250 women and about 1,600 men developed HPV-related carcinomas of the cervix, vagina, vulva, anus, penis, or oropharynx every year in Germany (24). HPV vaccination in Germany is recommended but still opportunistic, as there is not yet a comprehensive school-based or otherwise organized HPV vaccination program. The Robert Koch-Institute (RKI) monitors HPV vaccination rates in Germany. According to the RKI, only 31.3% of 15-year-old girls were fully vaccinated in 2015 (25), and HPV vaccination was far less well received than other childhood vaccines. Thus, the potential of cancer prevention through vaccination is not fully realized, and there is an urgent need to find entry points for improving HPV vaccination rates.

In the study PRÄZIS (prevention of cervical carcinoma and its precursors in women in Saarland), we analyzed the HPV vaccination situation by evaluating routine billing data for HPV vaccination and other health measures, such as preventive check-up examinations. Data were provided by the Association of Statutory Health Insurance Physicians Saarland (Kassenärztliche Vereinigung Saarland, KVS) and two statutory health insurance funds, AOK Rheinland-Pfalz/Saarland (AOK) and IKK Südwest (IKK). We selected Saarland as a model region, a federal state in western Germany with relatively low migration (26) and relatively high incidence rates for cervical, vulvar, and head and neck cancers in women (4). Our study provided new findings, particularly on the age-specific implementation of HPV vaccination by specific medical specialist groups and on the uptake of HPV vaccination compared with other preventive measures among girls and young women. We found that pediatric medical check-up examinations at the HPV vaccination age of 9-14 years, called “U11” and “J1”, can provide an excellent opportunity to deliver HPV vaccination in an age-appropriate manner. Specifically, our data show that there is a significant association between participation in U11 or J1 check-ups and HPV vaccination, and that the day of U11 has a high acceptance for HPV vaccination in the year of the U11 preventive check-up.

## Materials and Methods

### Ethical statement

In this study, we analyzed pseudonymized prospective and retrospective statutory health insurance billing data. The Ethics Committee of the Saarland Medical Association reviewed, approved all study procedures, and waived the requirement for informed consent (Ethikkommission Kenn-Nr. 186/17). All procedures were performed in accordance with the recommendations of the data protection authorities (Saarland University, Saarland Ministry of Social Affairs, Health, Women and Family Affairs, Saarland Independent Data Protection Center).

### Data sources

Pseudonymized routine data available from the KVS (2013-2019), IKK (2012-2019) and AOK (2009-2018) served as data sources. In addition to general information, service codes according to the uniform valuation standard (EBM) represented an essential component of the analyzed variables. Where necessary, the data from KVS and IKK were linked by automated record linkage in a study database and analyzed.

### Inclusion and exclusion criteria, baseline characteristics of the study population

The KVS and IKK datasets contained data on females who had received services from physicians in Saarland. This included individuals who resided outside of Saarland (S2 Fig). Saarland residents were selected by zip code. Missing data on gender were imputed if the gender could be inferred from more than 50% of the remaining data. The data sets and study populations used to answer the respective questions are listed in the corresponding figure and table legends. An overview of the data sets used and the operationalization of the variables for the individual questions is provided in Table 1.

**Table 1.**
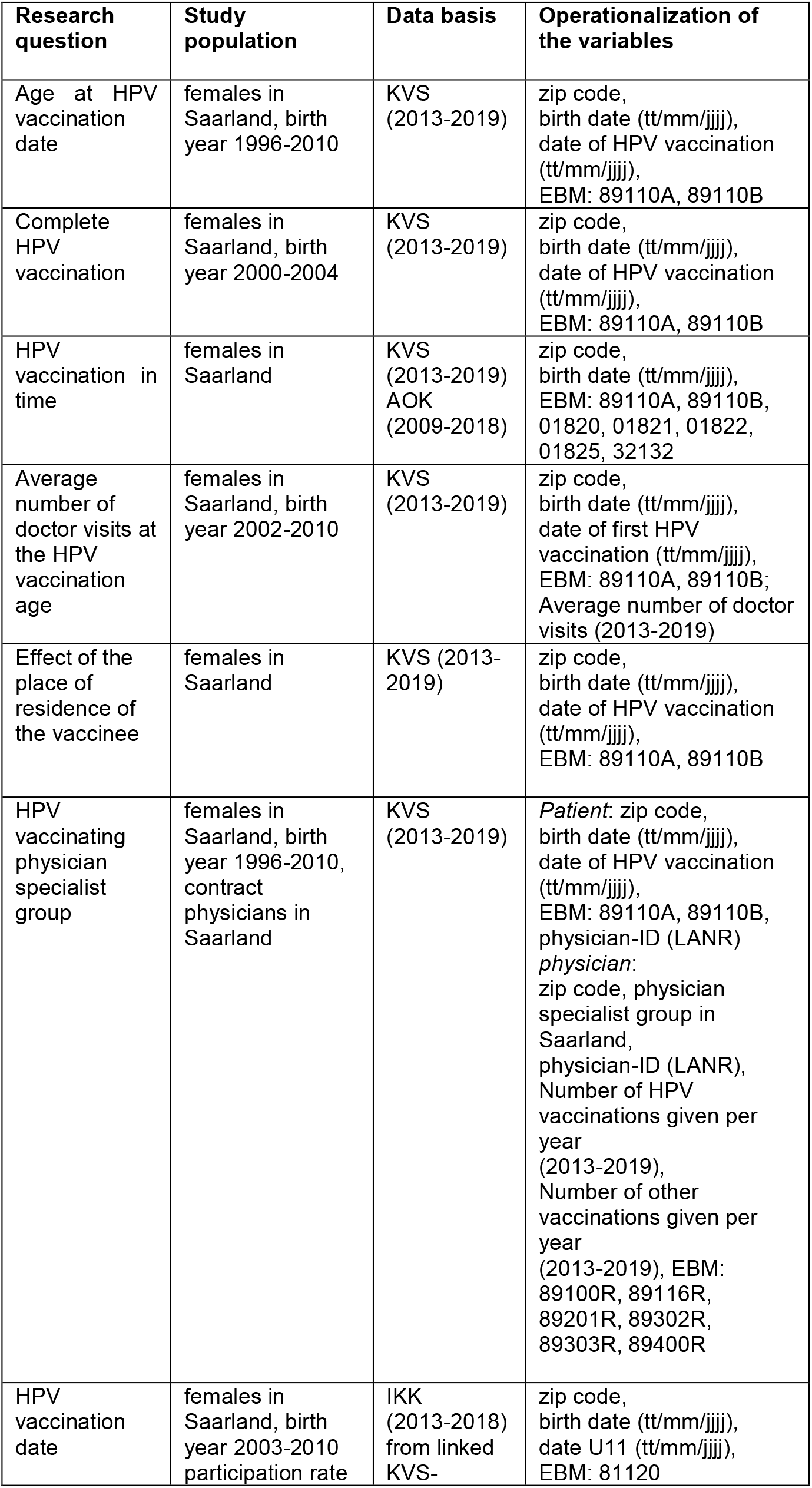
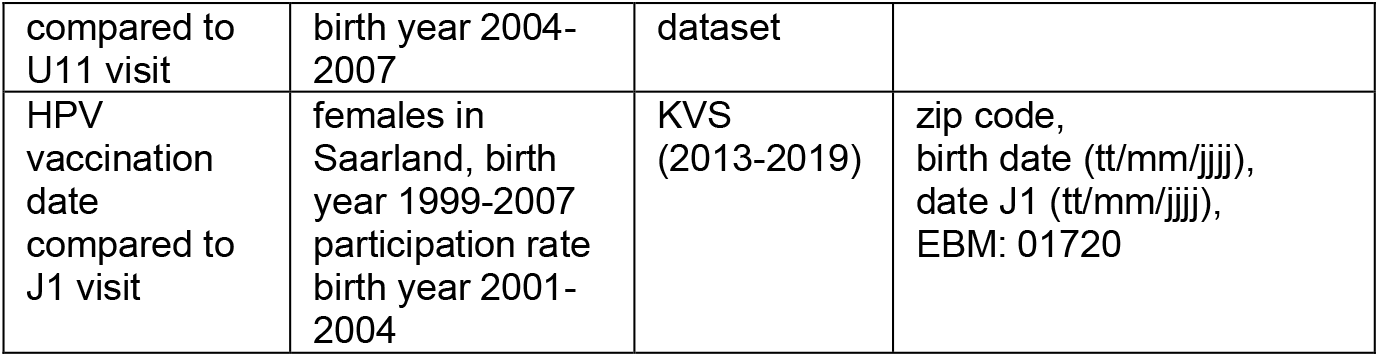
Overview of the data sets used and the operationalization of the variables for the individual research questions

### Technical procedure for pseudonymization

For all personal information, instead of transmitting identity data in plain text (last name, last name part 2, last name part 3 or additions, first name, first name part 2, first name part 3, maiden name, maiden name part 2, maiden name part 3, former name, former name part 2, Earlier Name Part 3, Date of Birth, Phonetically Standardized Last Name, Phonetically Standardized First Name, Phonetically Standardized Birth Name, Phonetically Standardized Earlier Name, Title, Title Part 2) only the corresponding hashed encrypted tokens derived were provided. Ciphering was performed using standardized plaintext features (one-way hash and symmetric IDEA encryption) by the KVS. This precluded recovery of the plaintext data. The linkage of the data was done using a record linkage procedure, which is routinely used in cancer registration and was adapted for the purposes of this study by the Saarland Cancer Registry (27).

### Technical procedure for data evaluation

The data analysis was based on the pseudonymized files in the study database. For this purpose, a raw database was created that complies with current data protection guidelines. Data security was ensured by an appropriate backup system. For evaluation purposes, a basic evaluation dataset was created in a separate instance, which emerged from the raw datasets through various data management procedures: data expansion, data and plausibility checks, variable calculations, recoding of variables. Subsequently, evaluation datasets were created from the basic evaluation dataset of the study database separately for each task within the scope of the work program on the research questions in a format compatible for the software analysis tools, which were only made available to the participating and authorized project partners.

In descriptive statistical analysis, absolute and relative frequencies were determined for qualitative data from the parameter list, such as HPV vaccination available, attendance of U8, U9, J1, etc., and means, medians, standard deviations, and ranks were calculated for quantitative variables, such as HPV vaccination age, etc. The data were additionally stratified by vaccination status: Once regarding vaccination present (yes/no) and the other according to vaccination status complete (yes/no). Statistical modeling was conducted using univariate and multiple regression analyses. Here, the potential influencing factors and predictors on the target variables HPV vaccination behavior, etc., that were present and relevant in the linked secondary data were examined. Logistic regression approaches were used, examining the following dependent and independent variables, among others: Dependent variable: HPV vaccination available (yes/no), Independent variables: Type of vaccinator, attendance at U8, U9, J1 examinations, age at vaccination, presence of other preventive measures. To account for possible multicollinearity, forward and backward variable selection according to the procedure of Wald was carried out.

Vaccination rates of 15-year-olds residing in Saarland from 2015 to 2019 were calculated based on the two EBM codes 89110A (first or subsequent vaccination) and 89110B (final vaccination). Considering the age at the start of the vaccination series and the recommended vaccination schedule (9–14-year-old girls: two doses at least five months apart, 15-year-olds: three doses), the completeness of the vaccination series was determined. The completeness of the vaccination series for 9–14-year-old girls at the start of the vaccination series was accordingly given, if a final dose was administered at the latest in the year of the 15th birthday or if two consecutive vaccination doses were administered at an interval of at least five months (two times 89110A). Completion of the vaccination series for 15-year-olds at the start of the vaccination series was given, if a final dose was administered no later than the year of the 15th birthday according to the billing code or if three consecutive vaccination doses were administered (three times 89110A). Absolute and relative frequencies were calculated. Analyses and data handling were performed using IBM-SPSS version 25 and R version 4. nQuery Advisor version 7.0 was used for calculations as part of the study design. Data were illustrated with Graph Pad Prism 9 (Graph Pad Software, San Diego, USA).

## Results

First, we determined the frequency of the first HPV vaccination in females in different age groups. For this purpose, we analyzed data from the KVS (years 2014 to 2019). Boys were not included in this analysis because there was no vaccination recommendation for boys at the beginning of the study.

After lowering the vaccination age from 12 to 9 years, we observed a clear increase in HPV vaccinations in 2015. The first HPV vaccine dose was increasingly given to younger girls aged 9 to 11 years (increase from 53 to 1,457 from year 2014 to 2019). The first HPV vaccination was more often given at the recommended age of vaccination (9-14 years) than catch-up vaccinations in 2015 to 15-17-year-old females (Fig 1A).

**Fig 1.**
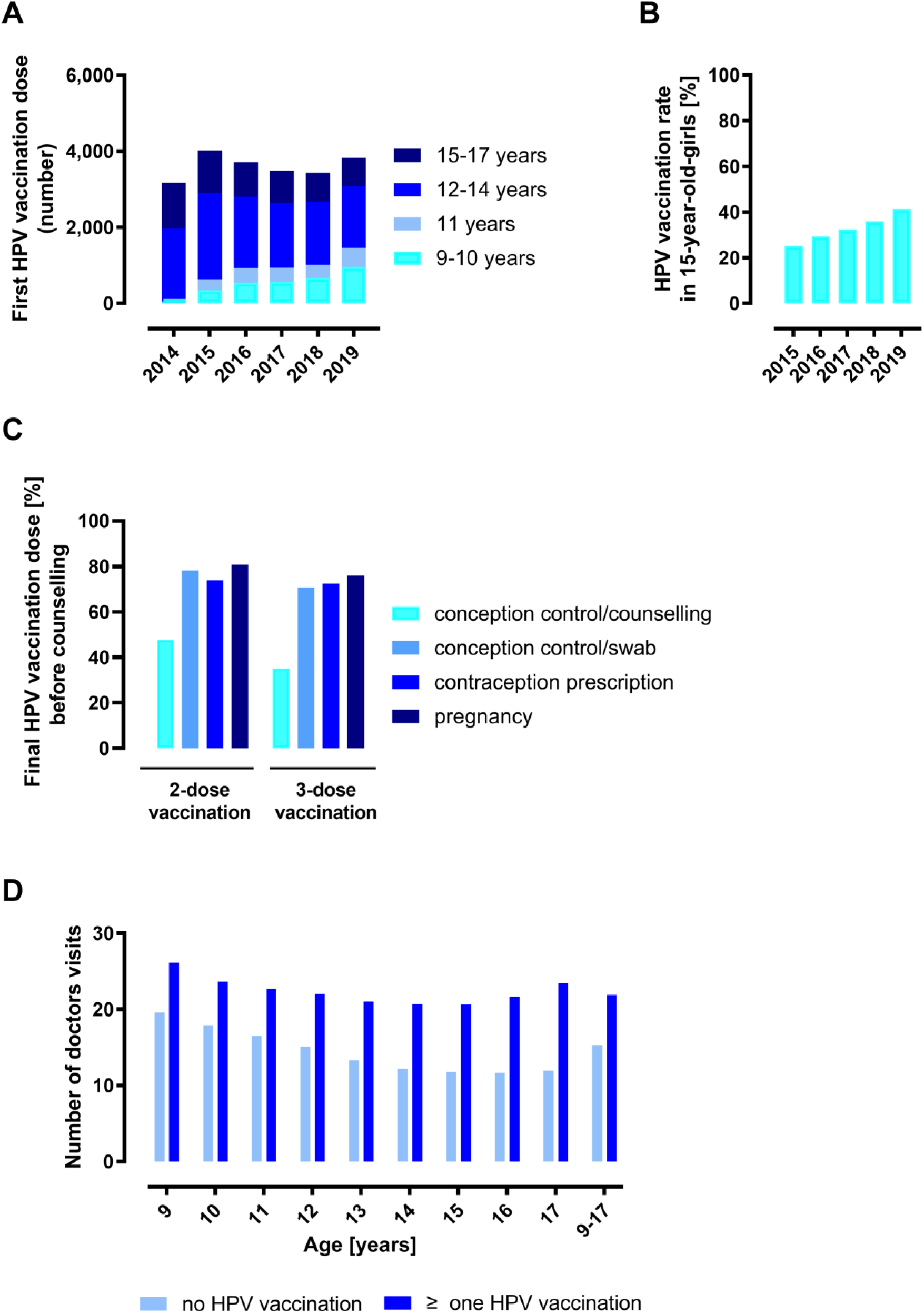
Numbers, rates, and timely finalisation of HPV vaccinations in females. (A) Numbers (absolute frequencies) of the first HPV vaccination by age and years of females in Saarland (KVS data 2013-2019). (B) Rates of fully vaccinated 15-year-old girls in Saarland. Percentage representation of the relative frequencies (data KVS 2015-2019). (C) Timely completion of HPV vaccination. Shown is the proportion of females with EBM code 89110B indicating the final HPV vaccination dose before or after the first date for counselling in the context of conception regulation (EBM code 01821, data KVS 2013-2019), smear collection in the context of conception regulation (EBM code 01825, data KVS 2013-2019), contraceptive prescription (EBM code 01820, data AOK 2009-2018), or pregnancy detection (EBM code 32132, data KVS 2013-2019). Percentage representation of relative frequencies. (D) Average number (absolute frequencies) of physician visits in billing years 2013 to 2019 for female individuals reaching the specified age in 2019 who received no HPV vaccination or at least one HPV vaccination (EBM code 89110A/B, KVS billing data, 2013-2019).

Data from the birth cohorts 2001 to 2004 living in Saarland were used to determine HPV vaccination rates for 2015 to 2019. The rates refer to girls who turned 15 in the corresponding year. For girls who received the first vaccine dose before their 15th birthday, a 2-dose schedule was sufficient for complete vaccination. This required the vaccine doses to be at least five months apart, according to the guidelines. For girls who received the first vaccine dose after their 15th birthday, a 3-dose schedule was necessary. For completeness, the required vaccine doses had to be administered no later than the end of the year in which the girls turned 15 years old. Our results show that the number of fully HPV-vaccinated 15-year-old girls in Saarland increased steadily between 2015 and 2019. While the HPV vaccination rate was 25.1% in 2015, it was 41.3% in 2019 (Fig 1B).

Subsequently, it was analyzed whether the last HPV vaccination took place before or after a date indicating the start of sexual intercourse, e.g., consultation or smear test in the context of conception regulation, prescription of contraceptives or indication of pregnancy. We would like to emphasize that this evaluation can only refer to those vaccinees for whom the relevant data were available in the analyzed period, i.e., who had undergone the relevant measures or were pregnant. In particular, in the case of prescription for a contraceptive, where data were only available from one health insurance company, or pregnancy, this was only a smaller fraction of those vaccinated (S1 Table). Fig 1C shows that in 47.82% (2-dose vaccination schedule) and 35% (3-dose vaccination schedule) of those vaccinated, the last HPV vaccine dose (as indicated by the EBM code 89110B) was administered before the time of contraceptive counseling, in 78.22% and 70.73%, before the time of contraceptive swab collection, in 73.85% and 72.35%, before the time of prescription for a contraceptive, and in 80.75% and 76.02%, respectively, before the first pregnancy.

We were also interested in the number of physician contacts at the vaccination age and whether there were differences between HPV-vaccinated and non-vaccinated females aged 9-17 years during the 2013-2019 data period. We found that HPV vaccinated girls had more physician contacts on average (20.68 to 26.18) than girls who were not vaccinated against HPV (11.67 to 19.64) (Fig 1D).

To further analyze access to HPV vaccination, we examined the regional distribution of medical specialist groups performing HPV vaccination. In particular, this included gynecologists, pediatricians, general practitioners, and internists, as well as the density of vaccinating doctors per municipality (per 10,000 inhabitants) in Saarland (Fig 2A). There were significant regional differences. In the district of the state capital Saarbrücken and in the vicinity of other larger cities such as St. Wendel, Saarlouis or Neunkirchen) with a higher density of physicians allowed to perform HPV vaccinations, all specialties were represented in a balanced way, whereas in more rural areas gynecologists and pediatricians were clearly less present.

**Fig 2.**
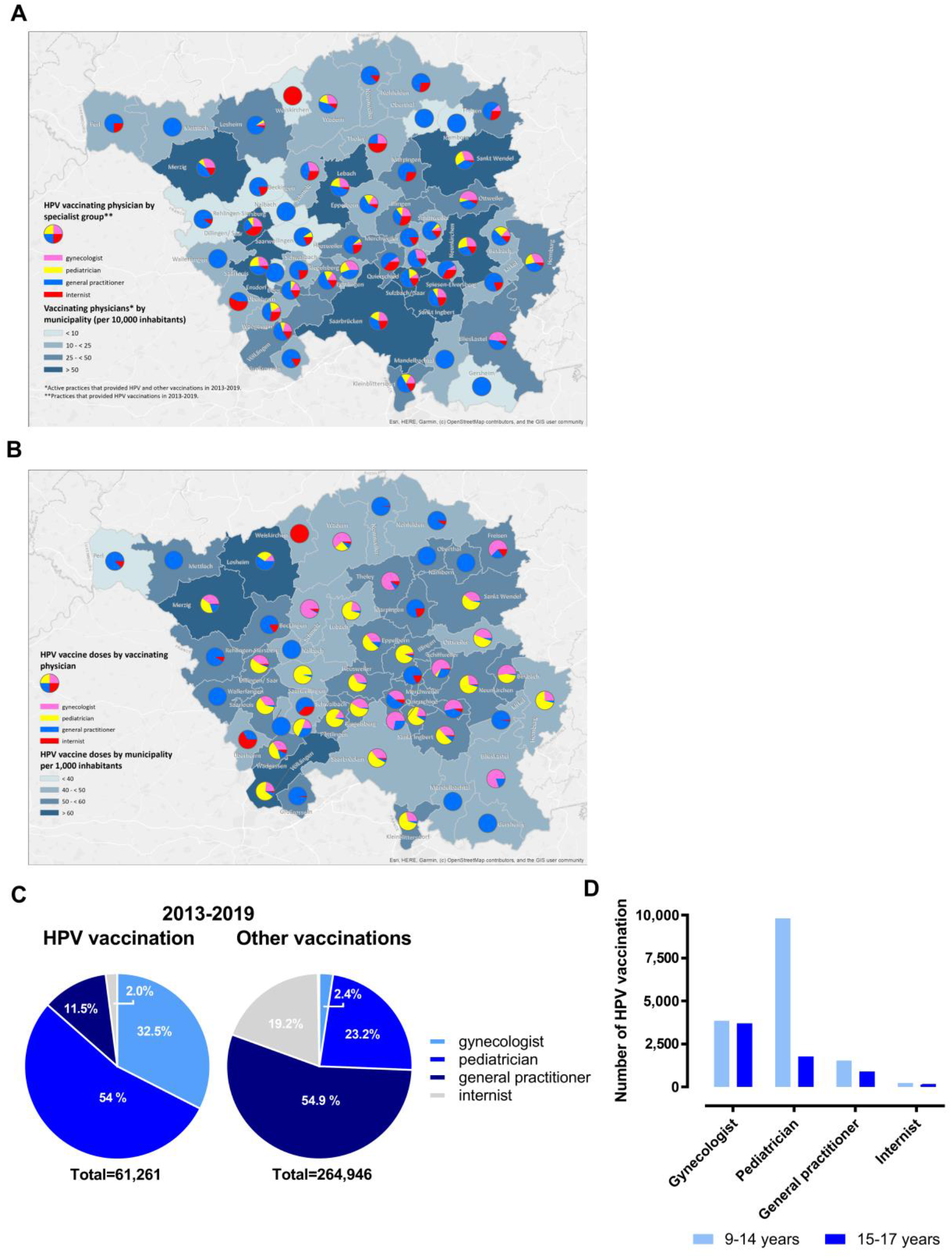
Relationship between HPV vaccination and vaccinating medical specialists. (A) Cartographic distribution of the vaccinating doctors by medical specialty (gynecologists, pink; pediatricians, yellow; general practitioners, blue; internists, red) based on Saarland municipality and per 10,000 inhabitants (data KVS 2013-2019). (B) Cartographic distribution of HPV vaccination doses by medical specialty and by Saarland municipalities, weighted according to the number of inhabitants (data KVS 2013-2019). (C) Administration of HPV vaccinations (total 61,261, left chart) or booster vaccinations (diphtheria, pertussis, tetanus, poliomyelitis, total 264,946, right chart) by different specialist groups. Percentage representation of relative frequencies in pie chart (data KVS 2013-2019). (D) Administration of HPV vaccine doses by medical specialty and age of vaccinees. Presentation of absolute frequencies (data KVS 2013-2019).

We then evaluated the spatial distribution of medical specialists with the respective administration of vaccine doses. The number of vaccine doses administered in the six districts showed little variation (S1 Fig). However, spatial analysis revealed significant regional differences in the administration of HPV vaccine doses per specialist in Saarland municipalities, weighted by population (Fig 2B). In 21 of 52 municipalities, pediatricians were the specialist group that administered the most HPV vaccine doses despite their lower absolute numbers (Fig 2A). In rural communities where pediatrician coverage was lower, HPV vaccine administration was compensated by other specialist groups, especially general practitioners, but also gynecologists.

To obtain a more general picture of vaccine administration in Saarland, we compared the relative contribution of different medical specialties to HPV vaccination and to booster vaccinations against pathogens other than HPV. Between 2013 and 2019, 54% of HPV vaccine doses were administered by pediatricians, 32.5% by gynecologists, 11.5% by general practitioners, and 2% by internists. In contrast, 54.9% of booster vaccinations against diphtheria, pertussis, tetanus, and poliomyelitis were administered by general practitioners and only 23.2% by pediatricians (Fig 2C). It should be noted, however, that the booster vaccinations evaluated in this study were not administered only to children, which may explain the significantly higher rate among general practitioners compared with pediatricians.

Analysis of HPV vaccination by discipline in terms of age of the vaccinees showed that 9-to 14-year-old girls were vaccinated mainly by pediatricians, whereas catch-up HPV-vaccinations of 15-to 17-year-old girls were performed mainly by gynecologists. These results underscore the importance of pediatricians in the successful delivery of HPV vaccination at the recommended age of 9-14 years.

The study also examined how HPV vaccination is delivered in relation to other preventive measures, such as childhood check-up examinations U8 (age 46-48 months), U9 (age 60-64 months), U11 (age 9-10 years) or J1 (age 12-14 years). In contrast to the nationwide implemented U8, U9 and J1, the U11 check-up is currently not offered by all statutory health insurance funds and is not billed in all cases by the KVS. Therefore, we were only able to evaluate the U11 participation rates for one health insurance company that provided the data. A major difference between the child preventive check-up programs in Saarland is a centrally organized invitation and reminder system for early U3 to U9, but not for later check-ups.

Our analysis shows that the numbers of U8 and U9 remained at a consistently high level between 2013 and 2019. In contrast, the number of J1 examinations at the HPV vaccination age of 12-14 years was significantly lower in 2013 and further declined steadily through 2019 (Fig 3A). Participation rates were determined for several birth cohorts in each case and ranged from 32% to 43% for U11 (Fig 3B) and from 25% to 31% for J1 (Fig 3C).

**Fig 3.**
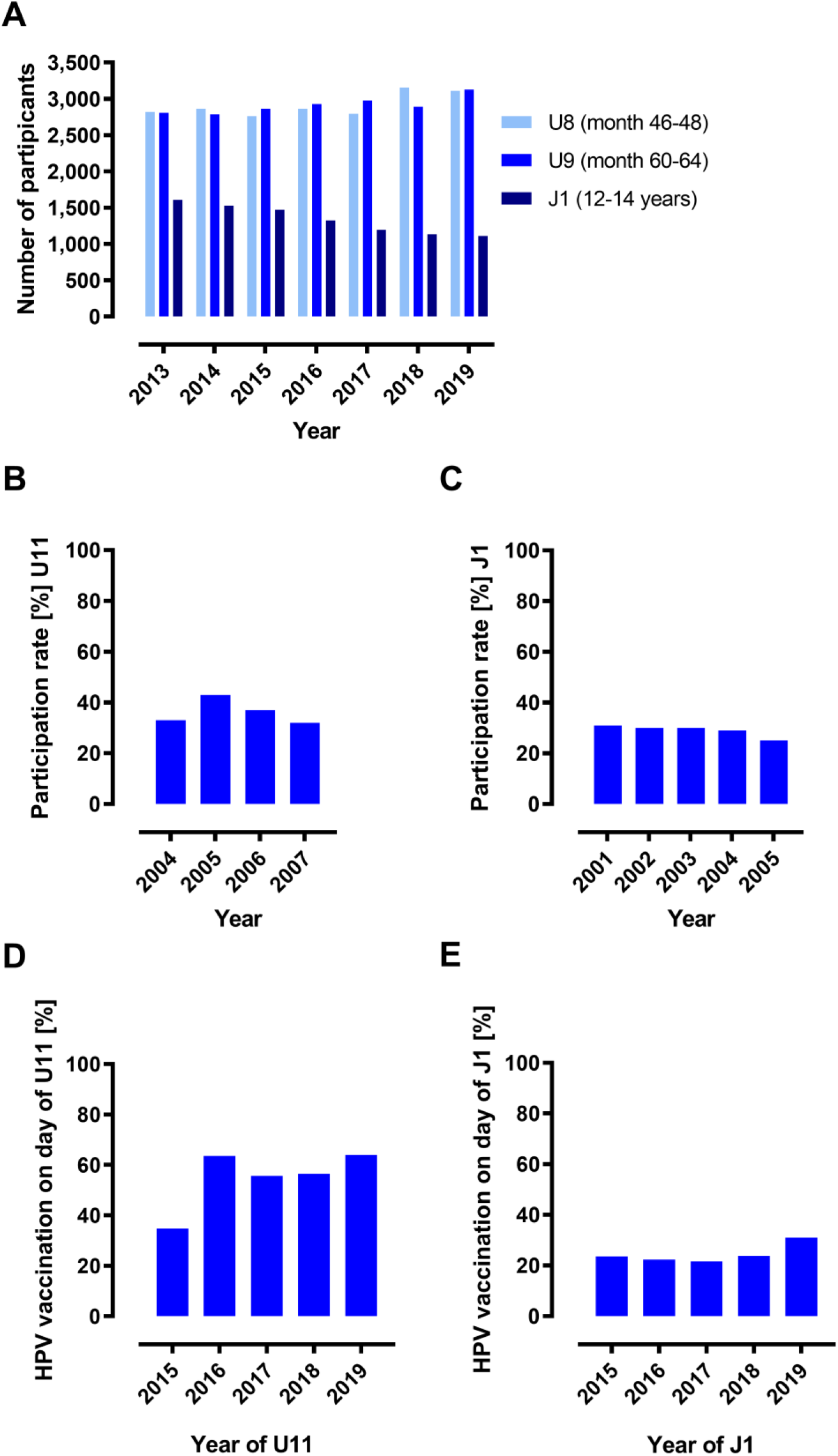
Relationship between HPV vaccination and participation in medical check-ups. (A) Number (absolute frequencies) of preventive medical check-ups among female individuals in Saarland (data KVS 2013-2019). U11 data are only shown from IKK health insurance (data were obtained from a linked IKK-KVS data set, 2013-2018). (B) U11 participation rates for girls of birth cohorts 2004-2007. Percentage representation of relative frequencies (obtained from a linked IKK-KVS data set, 2013-2018). (C) J1 Participation rates for female individuals of birth cohorts 2001-2004. Percentage representation of relative frequencies (data KVS 2013-2019). Utilisation of the U11 (D) or J1 (E) appointment for HPV vaccination by females in the year of U11 or J1, respectively. Percentage representation of relative frequencies. Data in (D) were obtained from a linked IKK-KVS data set, 2013-2018; data in (E) from KVS 2013-2019.

Logistic regression analysis was performed to examine the association between HPV vaccination and U11 and J1 participation during the HPV vaccination age of 9-14 years. Regression analyses for the birth cohorts 2004 and 2005 found that girls who participated in U11 check-up had 5.3 times (95% CI: 4.66 - 6.11) higher odds of receiving HPV vaccination than girls who did not attend the U11 (p <0.001). Girls who participated in J1 were 10.03 times (95% CI: 8.96-11.22) more likely to receive HPV vaccination than girls who did not attend J1 (p <0.001). In the year of the individual U11 examination, up to 63.9% of those vaccinated were vaccinated against HPV on the day of the U11 examination (Fig 3D). In the year of the individual J1 check-up, up to 31% of those vaccinated against HPV were vaccinated on the day of the J1 examination (Fig 3E). The analysis did not rule out the possibility that girls were vaccinated in an earlier year.

The U11 appointment therefore seems particularly appropriate for early HPV vaccination at the age of 9-10 years.

## Discussion

Our study based on routine health insurance data not only provides robust data on HPV vaccination in a German model region in recent years but also provides new starting points for improving HPV vaccination rates.

We show that HPV vaccination rates in Saarland among 15-year-old girls in 2019 are still well below 50%, far below vaccination rates in other countries such as Australia (28) or in other European regions, e.g., UK (29). High HPV vaccination success rates in other countries may be based on the early implementation of organized HPV vaccination programs, such as school-based vaccination in United Kingdom, Canada and Australia (30). In Germany HPV vaccination is not yet organized but rather opportunistic, although it was called for early after HPV vaccination was introduced (31).

Our data show that overall, HPV vaccination rates have steadily increased since 2015 in Saarland. This was particularly due to an increase in the proportion of 9-to 11-year-old girls among HPV vaccinated. Lowering of the HPV vaccination to age 9 with the 2-dose schedule (22) resulted in more HPV vaccinations completed. This corresponds to the increase in HPV vaccination rates until 2018 recently shown by (32, 33), although the exact rates might differ due to differences in the databases (we have limited our evaluations to Saarland resident females) or possibly also calculation modalities.

Prophylactic HPV vaccination should be completed ideally before first sexual intercourse (21), since HPV incidence peaks shortly after sexual debut (34). Therefore, we examined the timing of completion in those who showed signs of early onset of intercourse. In our study the final HPV vaccination was 35 to 80.75% after the date of a counseling session or first date of smear collection in the context of conception management, the prescription of contraceptives, or pregnancy. This assessment could only refer to those who had undergone the relevant measures or were pregnant. Although the groups studied represent only fractions of those vaccinated, our data suggest that, particularly in these vulnerable subpopulations of females, vaccination was not completed in a timely manner on a substantial scale.

Interestingly, there was no pronounced urban-rural difference in HPV vaccine doses administered in Saarland. In contrast, the specialty of the vaccinating physician played an important role. Pediatricians administered the most HPV vaccine doses and a particularly high proportion of vaccinations to girls of the recommended vaccination age between 9 and 14 years. Gynecologists, on the other hand, administered more HPV catch-up vaccinations than pediatricians. In contrast, booster vaccinations against other diseases were more likely to be administered by general practitioners. In Saarland, in communities where there are fewer pediatricians, general practitioners and gynecologists increasingly administered HPV vaccinations. This suggests that all three disciplines, pediatricians, gynecologists, and general practitioners, play an important role in different areas or vaccinee ages and should be involved in HPV vaccination campaigns to increase vaccination rates in the future.

In general, the adherence of vaccinated individuals to their physicians appears to play an important role. HPV-vaccinated individuals had significantly more physician contacts during the vaccination age than non-vaccinated individuals. Each visit to the doctor is therefore a valuable opportunity to be vaccinated against HPV.

Visiting the pediatrician is especially important to get vaccinated as early as possible. In Germany, children and adolescents who are not yet of vaccination age are invited and reminded to attend the preventive check-ups U3-U9. These early visits are therefore consistently well used. Child and adolescent health check-ups are also offered at the recommended immunization age of 9-14 years. The J1 examination is conducted nationwide for 12–14-year-olds, but in Saarland without an invitation system. In contrast to the U8 or U9 examinations, participation in the J1 examinations was significantly lower and has continued to decline over the years. The U11 is intended for the even younger ones at the age of 9-10 years. However, it is currently not offered and financed by all health insurance funds in Germany, and, even if financed, is not always billed through KV. For U11, the Saarland-wide participation rate could therefore not be determined, because billing data from only one Saarland health insurance company (IKK) were available for evaluation. However, our analysis of the available data showed that there is a significant correlation between the participation in the U11 or J1 check-up examinations and HPV vaccination. Those who attend the corresponding pediatric or adolescent medical examination have a significantly higher chance of being vaccinated against HPV. A positive association is already known from a study on J1 examination and HPV vaccination (35). Other studies could only show a weak or no correlation with participation in a J1 examination (36).

Our data also show that the day of U11 has a high uptake for HPV vaccination in the year of the U11 examination. In our experience, parents at this age are more likely to accompany their children to the screening appointment than to J1, so parental consent for vaccination can be obtained on the spot and thus vaccination can take place on the same day. The prerequisite for this is that the HPV vaccine is stocked in the doctor’s office, which is the case in Saarland, and should be rolled out nationwide. Organized combined invitation to U11/J1 and HPV vaccination could address issues of different population subgroups and thus reach individuals of different geographical regions, ethnities, socioeconomic status and religions. In addition, HPV vaccination in younger individuals is seen as a “normal childhood vaccination” rather than an “STI- or cancer vaccine” and can be decoupled from the issue of sexual activity at a young age.

In summary, our data suggest that the integration of an organized HPV vaccination program into U11 and J1 health check-ups could be very well suited to increase vaccination rates in Germany. This could be an alternative to school-based vaccination, which is not yet implemented in Germany. In particular, the U11 date is the ideal time for an intervention to increase HPV vaccination rates in Germany. This is in line with a strong recommendation of the S3 guideline “Vaccination prevention of HPV-associated neoplasms” (37) to improve HPV vaccination rates, for which our study now provides the first evidence.

Our data are based on statutory health insurance billing data and are therefore more robust than data collected via questionnaires. However, our study also has limitations. It should be mentioned that 10-15% of people in Germany have private insurance and were therefore not included in our study. Their vaccination behavior may differ from that of those with statutory insurance. Furthermore, our project started before 2018, when there was no vaccination recommendation for boys in Germany. Vaccination of boys may, however, ensure more robust herd protection. Both aspects, privately insured children including boys, should therefore be considered in a future intervention study to test the effect of organized HPV vaccination.

## Data Availability

All relevant data are within the manuscript and its Supporting Information files.

## Acknowledgements

The authors thank Adam Bekhit for technical support of this study.

## Supporting Information

**S1 Table:** Timely completion of HPV vaccination. Shown are the numbers of females with EBM code 89110B before or after the first date for counselling in the context of conception regulation (EBM code 01821, data KVS 2013-2019), smear collection in the context of conception regulation (EBM code 01825, data KVS 2013-2019), contraceptive prescription (EBM code 01820, data AOK 2009-2018), or pregnancy detection (EBM code 32132, data KVS 2013-2019).

|  | Final HPV vaccination | Medical indication for doctors' visits |  |  |  |
| --- | --- | --- | --- | --- | --- |
|  |  | Conception control/counselling | Conception control/swab | Contraception prescription | Pregnancy |
| **2-dose vaccination** | Before | 2,172 | 1,681 | 257 | 541 |
|  | After | 2,370 | 468 | 91 | 129 |
| **3-dose vaccination** | Before | 2,287 | 2,938 | 806 | 929 |
|  | After | 4,247 | 1,216 | 308 | 293 |

**S1 Fig.**
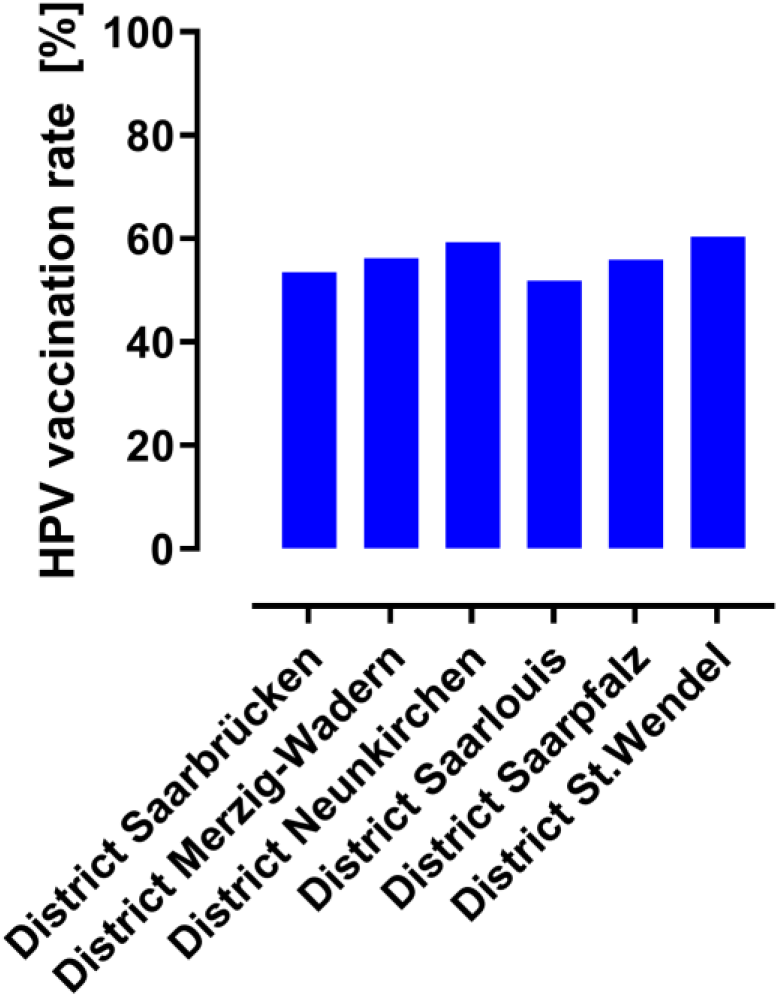
Proportion of patients from a district in Saarland who have received at least one HPV vaccination (percentage representation of the relative frequencies, data KVS, 2013-2019).

**S2 Fig.**
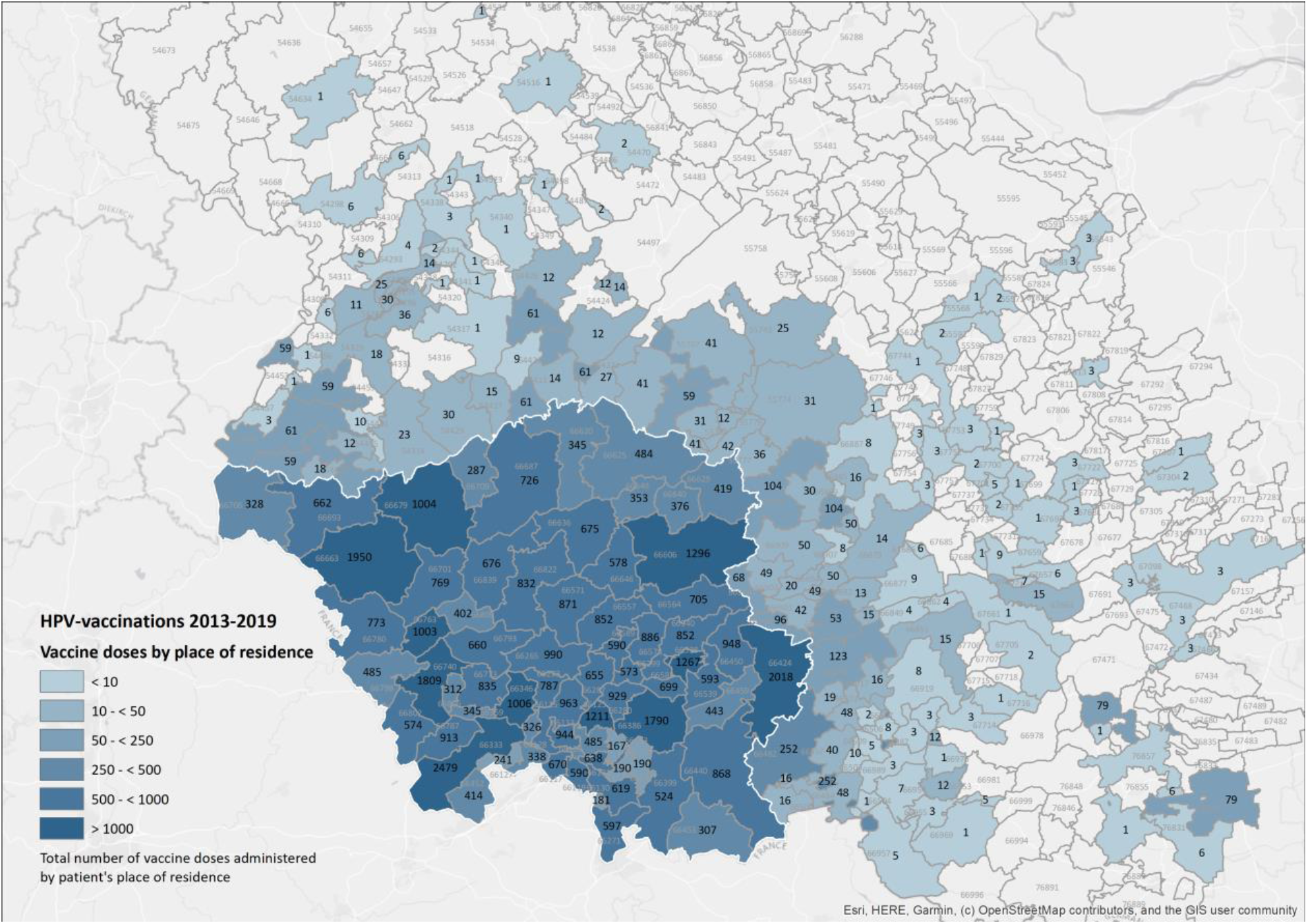
Cartographic representation of the cumulative HPV vaccination doses billed by the KVS between 2013-2019, administered to females resident in Saarland and neighbouring regions.

